# Urban Wastewater as Environmental Reservoirs of Multidrug-resistance *Enterobacteriaceae* during the COVID-19 Pandemic in India

**DOI:** 10.1101/2025.01.01.24319799

**Authors:** Hina Qamar, Touseef Hussain, SaadBin Javed

**Author notes:** shared equal authorship (Hina Qamar, Touseef Hussain).

## Abstract

Secondary bacterial infections have been associated with COVID-19, leading to increased antibiotic consumption and exacerbating the phenomenon of antimicrobial resistance (AMR). The present study evaluated the prevalence of multidrug-resistant (MDR) *Enterobacteriaceae* (specifically *Escherichia coli* and *Klebsiella pneumoniae*) in wastewater collected from three different Western Uttar Pradesh (WUP) zones during the pandemic. A total of 150 isolates were identified both biochemically and using VITEK-2. Antibiotic susceptibility testing was achieved using the disc diffusion method. Of the 150 isolates, *Escherichia Coli (E. coli)* (62%) was the predominant bacterial species, followed by *Klebsiella pneumoniae (K. pneumoniae)* (24%), other *Enterobacteriaceae* (8%) and a mixed population of bacterial isolates (6%). Also, 74% of the isolates were resistant to one or more antibiotic classes. A high percentage of MDR was detected in *E. coli* (57.14%), followed by *K. pneumoniae* (28.57%). However, compared to *E. coli*, the MDR prevalence of *K. pneumoniae* in different zones was significant (*p*-value=0.0081). Of the total isolates, 79.3% were extended-spectrum beta-lactamases (ESBL)-producers. The resistance to ESBL in *E. coli* was higher than in *K. pneumoniae*. The highest resistance was observed against penicillin, cephalosporin, quinolone, and fluoroquinolone. Notably, two *K. pneumoniae* isolates and *Pantoea agglomerans* showed resistance to all antibiotics tested, including colistin. To our knowledge, this is the first study performed in the Indian WUP region during COVID-19. Resistance to colistin and the multidrug resistance observed in the current study is a cause for concern due to the constant exposure of people to polluted waters in India, especially within resource-limited environments. The study calls for further studies to understand the full extent of the problem and implement mitigation measures.

## 1. Introduction

The recent coronavirus disease (COVID-19) pandemic has claimed millions of lives worldwide and remains a public health challenge. COVID-19 is primarily a respiratory infection, although the probability of secondary infections after COVID-19 has been reported in some patients [1,2]. This increased antibiotic use and antimicrobial resistance (AMR) [3]. AMR is among the most serious threats to global health and the economy [4,5]. Lack of awareness, overcrowding, co-infections, insufficient diagnostics, poor health infrastructure and financial compensation of doctors exacerbate AMR in developing countries such as India [6].

In the first pandemic wave, medical misadventures worldwide led to patients’ indiscriminate use of antibiotics and other medicines. Non-regulatory use of antibiotics in India constitutes about 90% of antibiotic sales prescribed by 70% of private healthcare practitioners. The findings revealed that COVID-19 guidelines were not followed during the pandemic, and about 216.4 million additional antibiotic doses were consumed to treat mild to moderate-severe acute respiratory syndrome coronavirus 2 (SARS-CoV-2) infections [7]. For example, 16.29 billion antibiotics were sold during the year 2020 in India alone. Antibiotic consumption in adults increased from 72.6% in 2018 and 72.5% in 2019 to 76.8% in 2020 [8]. Furthermore, the shortage of healthcare professionals and doctors in India and people’s fear of the pandemic fueled their interest in exploring Google search trends for self-prescription [6]. Antimicrobials were used blindly in hospitals, clinics, and Intensive Care Units (ICUs) to prevent co-infections and the SARS-CoV-2 viral infection and skyrocketed during the second pandemic wave [6]. Studies have reported that *Enterobacteriales* and *Staphylococcus aureus* are the most common pathogens causing these secondary infections [9]. Thus, the indiscriminate use of antibiotics could lead to AMR in these pathogens.

The World Health Organization (WHO) has published a list of priority pathogens. Of the 12 families listed by WHO, *Enterobacteriales* pose the greatest threat to human health, especially when transmitted throughout the environment [10]. *Enterobacteriales* constitute gut microbiota; most are primary nosocomial and community-acquired infection-associated pathogens [11]. *E. coli* and *K. pneumoniae* are common pathogens of this family in which AMR has escalated in recent decades [11]. Carbapenem-resistant and 3rd generation cephalosporin-resistant *Enterobacteriaceae* (CRE) are at the highest priority on the WHO list of pathogens [10]. Resistance to carbapenems, β-lactams, methicillin, vancomycin, fluoroquinolones and ampicillin are among the resistance threats on the WHO priority pathogen list [10,12]. Both *E. coli* and *K. pneumoniae* are common intestinal bacteria. Most *E.coli* strains are harmless, but some cause severe foodborne illnesses [13–16]. On the other hand, *K. pneumoniae* causes life-threatening infections [17]. Fluoroquinolone resistance in *E. coli* and carbapenem resistance in *K. pneumoniae* have spread worldwide. Such spread limits therapeutic options making many infections incurable even with existing antimicrobials [11]. Colistin is the last option for severe infections caused by carbapenem-resistant *Enterobacteriaceae* strains, although resistance to it has also been reported in various parts of the world [18–21]. However, no effective antibiotic treatment is currently available for colistin resistance [21,22].

The residual un-metabolised antibiotics used in humans and animals are excreted in faeces and enter sewage systems, intensifying AMR selection pressure [23]. However, in developing countries such as India, wastewater treatment plants (WWTPs) are usually non-existent or are not well equipped to remove these residual antibiotics and antibiotic-resistant bacteria, which usually find their way into the environment (especially aquatic ecosystems) and increase global AMR prevalence [24–26]. Thus, wastewater, having ubiquitous microbiota, has become a hotbed of AMR transmission.

Based on these premises, the present study focused on isolating antibiotic-resistant-*Enterobacteriaceae* from wastewater during the COVID-19 pandemic in the WUP region of India. Particular attention was placed on MDR *E.coli* and *K. pneumoniae* (carbapenem-resistant and 3^rd^-generation cephalosporin, e.g., extended-spectrum β-lactamases, ESBL). The study further investigated colistin-resistant *Enterobacteriaceae* due to the recent concern in colistin resistance. The aim of the study is to draw the attention of government, policymakers and the scientific community to the spread of MDR in non-metropolitan cities so that the infrastructure facilities in other cities of India could be upgraded. It would be expected that the study’s findings will provide valuable information on the potential threat to which populations could be exposed when using water from sources polluted with these resistant bacteria.

## 2. Materials and Methods

### 2.1 Description of study site

Western Uttar Pradesh region covers the western Uttar Pradesh districts, with distinct demographic, economic and cultural patterns. This region is the fastest growing in the area and has a population of over 71 million inhabitants, most of whom are engaged in agriculture. Uttar Pradesh experiences high social disparity, characterized by a higher population living in areas without pipe-borne water and proper sanitation facilities. Thus, many people depend on private tube wells, pumps and latrines. A full description of the region has previously been published [48].

### 2.2 Sample collection and enrichment

Fifteen wastewater sampleswere collected in triplicates between 2020 and 2021 from untreated drainage systems in different zones in WUP, covering both the metropolitan and non-metropolitan cities of National Capital Regions (NCR) (Zone 1, covering Ghaziabad, Noida, and Sahibabad areas; Zone 2, covering Agra, Hathras, and Aligarh areas; and Zone 3 covering Meerut, Bulandshahar and Bijnor areas). Each sample was collected in sterile conical flasks and store at 4°C. Triplicate samples from each sampling site were pooled per sampling round. Thus, 100 ml of each sample was filtered through a 0.45 μm filter, followed by enrichment of the filters in Enterobacteria Enrichment Broth-Mossel (Hi-Media Laboratories Pvt. Ltd, Karnataka, India) to isolate different Gram-negative *Enterobacteriaceae* bacterial strains.

### 2.3 Isolation, purification and characterisation of bacterial isolates

Samples from each enriched culture were inoculated separately onto MacConkey Agar (MCA) (Hi-Media Laboratories Pvt. Ltd, Karnataka, India) in duplicates to obtain *E. coli* and *K. pneumoniae* strains. The plates were incubated at 37 °C for 24 h and then observed for bacterial growth. All isolates with differential colony morphology were chosen for a specific plate. After that, the isolates were purified by repeatedly subculturing on MCA, in order to obtain pure colonies, and phenotypically characterized via Gram staining, microscopy, and standard biochemical reactions. For the initial screening of the isolates, biochemical characterization was performed to understand the strain. Some purified bacterial colonies were picked separately from each plate and subsequently suspended in 3 ml of normal saline to form a homogenous suspension of 0.5 McFarland standards (≥0.1 OD at 620 nm wavelength). Afterwards, biochemical tests were performed using the KB003 TM Hi25 *Enterobacteriaceae* Identification Kit (Hi-Media Laboratories Pvt. Ltd, Karnataka, India) according to the instructions provided with the kit. Later, biochemically characterised isolates were further confirmed by identification through VITEK automated system (bioMérieux, Nürtingen, Germany).

### 2.4 Antibiotic Susceptibility Testing

For all bacterial isolates, sensitivity to appropriate antibiotics was determined by the standard disc diffusion method (Bauer et al., 1966) with antibiotic discs (Hi-Media Laboratories Pvt. Ltd, Karnataka, India) (TableS1, Supplementary Materials). The results were interpreted per the Clinical and Laboratory Standards Institute (CLSI) guidelines (CLSI, 2020).

### 2.5 Detection of ESBL

Possible ESBL-producing isolates were screened using the antibiotics ceftriaxone (30 µg) and cefoperazone (75 µg). Isolates with < 25 mm zone of inhibition for ceftriaxone and <22 mm for cefoperazone were considered ESBL-positive. In addition, the potentiation of cefoperazone activity in the presence of cefoperazone+sulbactam was confirmed. An increase in diameter of 5 mm was considered positive for ESBL [50]. According to Centers of Disease Control and Prevention (CDCs definition, https://www.cdc.gov/infectioncontrol/guidelines/mdro/index.html), the isolated strains were considered multidrug-resistant (MDR) when resisting to more than one or more classes of antimicrobial agents.

### 2.6 VITEK identification and determination of multidrug resistance

Biochemically characterised bacterial isolates were further identified using VITEK2 (bioMérieux, Nürtingen, Germany) as per the manufacturer’s manual. Testing was done with VITEK^®^ 2 PC LIS-compatible software using ID-GN VITEK^®^cards. Antibiotic susceptibility validation was carried out using AST-N280 cards for pure isolates. The results were interpreted based on the minimum inhibitory concentrations (MICs) as per the CLSI standard breakpoints(CLSI, 2020). The 2^nd^ and 3^rd^-generation cephalosporins (ceftazidime, cefotaxime, and cefuroxime) were used alone or in combination with clavulanic acid. For ESBL validation, a reduction of growth in the presence of clavulanic acid was considered indicative of ESBL production. In addition, colistin-positive isolates were further screened to confirm the minimal inhibitory concentrations (MICs) using the colistin broth disk elution (CBDE) method (Simmer et al., 2019). Briefly, a single colony was taken and adjusted to 0.5 McFarland standards in 0.9 % NaCl. About 50 μL of the suspension was diluted in 11.5 mL of Mueller-Hinton II broth (Oxoid, Wesel, Germany). The broth was transferred to 96-well antimicrobial susceptibility testing plates, followed by incubation at 37 °C for 24 h and checked for turbidity at the bottom of the well (CLSI, 2020).

### 2.7 Statistical analysis

Data were entered into Excel (Microsoft Office, Microsoft Corporation, Redmond, WS, USA) and analyzed using STATA16 statistical software (StatCorp., Austin, TX, USA). Qualitative variables were described with absolute and relative frequencies. Associations between categorical variables were tested with chi-square. Differences be-tween proportions were tested with the z-test. Quantitative variables were represented by measures of position and variability. Differences between means were tested with the Student’s t-test distribution for independent samples.

## 3. Results

### 3.1 Isolation, Purification and Characterisation of Bacterial colonies

Between 2020 and 2021, a total of 150 isolates belonging to the family *Enterobacteriaceae* were isolated and screened from untreated wastewaters in three different zones of the WUP region, covering both metropolitan and non-metropolitan cities. Phenotypically, these isolates were identified as presumptive *E. coli* and *K. pneumoniae*.

Further characterisation of these isolates identified them as *E. coli* (93/150; 62%), *K. pneumoniae*(36/150; 24%), other *Enterobacteriaceae* (12/150; 8%, *e.g., Pantoea agglomerans*, *Alcaligenes faecalis* and *Shigella*) and mixed population (9/150; 6%, unidentified) (Figure 1).

**Figure 1.**
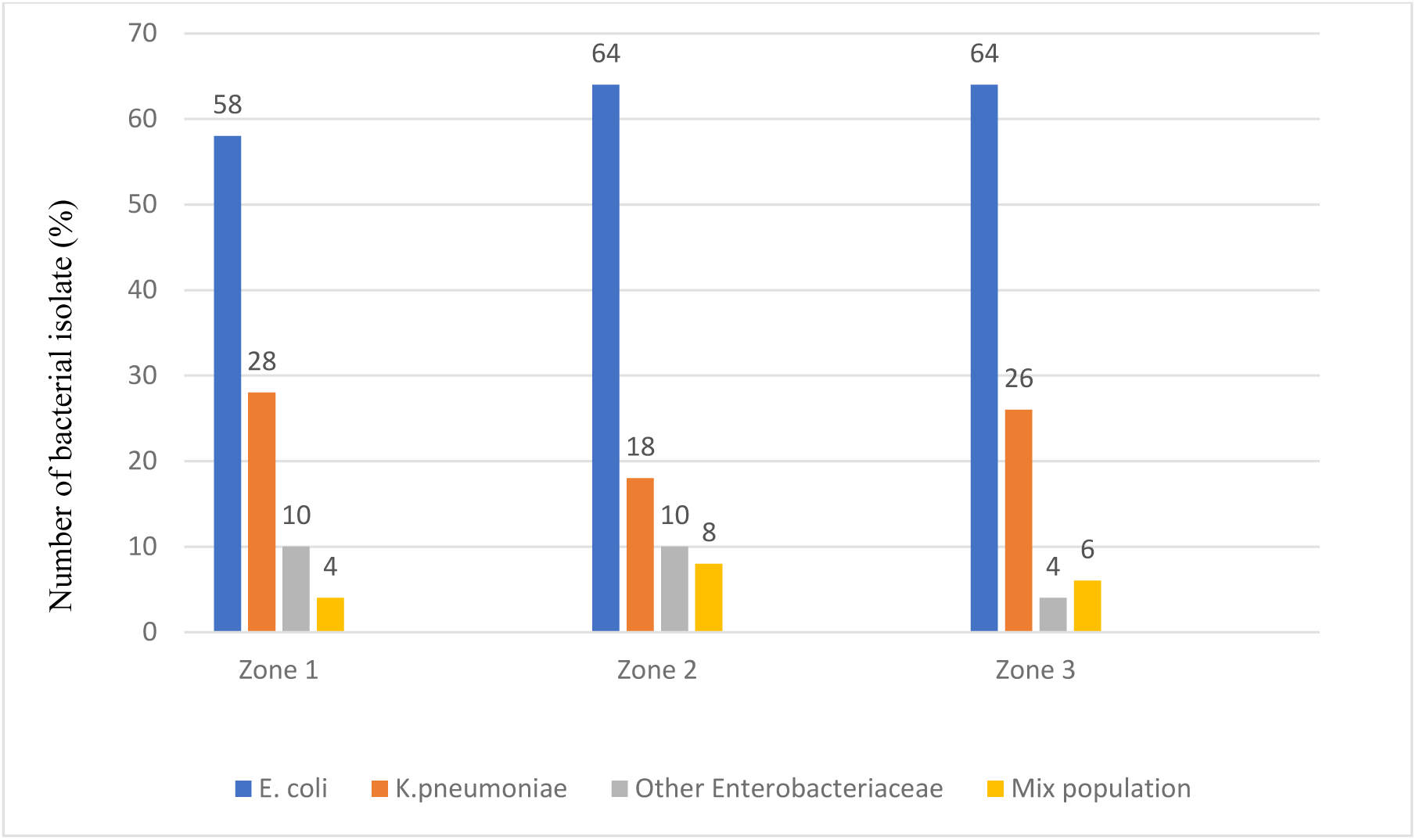
Bacterial isolates in different zones of Uttar Pradesh region, India.

### 3.2 Antimicrobial susceptibility

The antimicrobial susceptibility of different *Enterobacteriaceae* isolates (*n* = 150) was tested against 19 antibiotics classified into eight classes reported in figure 2. Of the isolates, 39 (26%) were susceptible to all antibiotics tested (except ampicillin), while 111 (74%) isolates showed resistance to one or more antibiotics. Out of the 111 resistant isolates, 88 isolates were identified as ESBL producers (79.3%). Notably, *E. coli* (57.14%), *K. pneumoniae* (28.57%) and *Pantoea agglomerans* (5.71%) exhibited multidrug resistance. The regional (zone-wise) antimicrobial susceptibility distribution is shown in Figure. 2.

**Figure 2.**
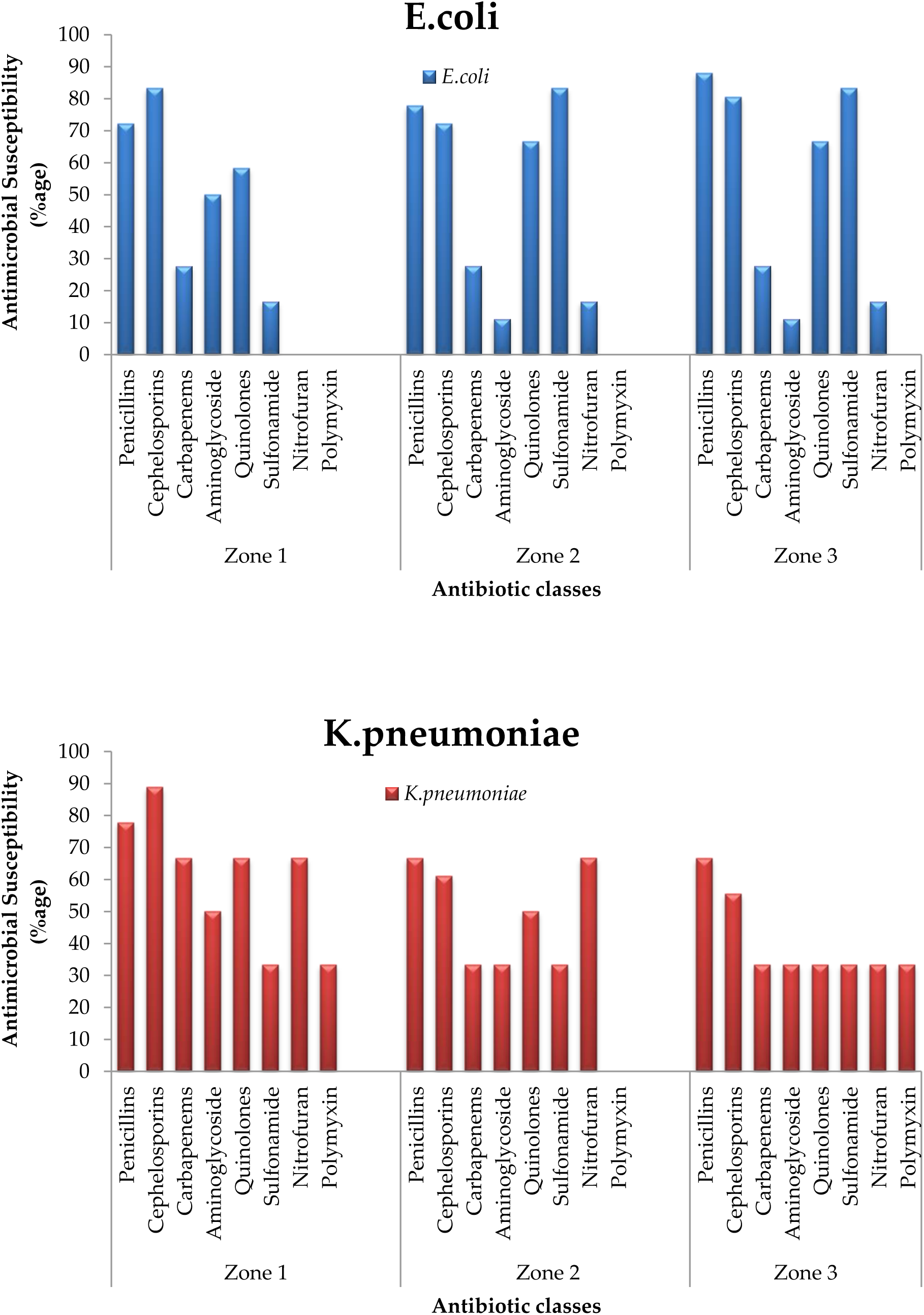
Antimicrobial susceptibility of different *Enterobacteriaceae* isolates obtained from three different zones.

As per figure 2, the prevalence of ESBL resistance for *E. coli* was higher in Zone 3 than the other zones. In contrast, it was higher for *K. pneumoniae* in Zone 1. In zone 1, for both *E. coli* and *K. pneumoniae*, cephalosporin resistance was higher than in zones 2 and 3. Furthermore, *K. pneumonia* had the highest car-bapenem resistance in zone 1. The highest resistance to aminoglycosides was observed in zone 1 for both *E. coli* and *K. pneumoniae*. For quinolones, *E. coli* showed the highest resistance in zones 2 and 3, while *K. pneumoniae* showed the highest resistance in zone 1. In addition, *E. coli* showed the highest resistance to sulfonamide in zones 2 and 3, while *K. pneumoniae* showed an almost identical resistance pattern in all three zones. For nitrofuran, the highest percentage of resistance was observed in *K. pneumoniae* in zones 1 and 2. Resistance to polymyxin was not observed in *E. coli*, while zone 1 and 2 isolates showed resistance to colistin. In general, Zone 1 had the highest multidrug resistance percentage. Also, MDR was almost identical in Zones 2 and 3, with slight variations. Multidrug-resistant *K. pneumoniae* populations were highest in Zone 1. On the other hand, Zone 3 had the highest population of MDR *E. coli*.

The percent MDR pattern in decreasing order was as follows: Zone 1> Zone 2> Zone 3. However, zonal total MDR statistical analysis showed a *p*-value=0.85 which is not significant. Beside, a significant resistance pattern for different antibiotic classes was observed for *E. coli* in all zones (Zone 1: *p*-value <0.0001; Zone 2: *p*-value=0.0124; Zone 3: *p*-value=0.0004). On the other hand, for *K. pneumoniae* different resistance pattern exists in different zones (Zone 1: *p*-value=0.0427; Zone 2: *p*-value=0.0415; Zone 3: *p*-value =0.2592). Nevertheless, in comparison to *E.coli* the MDR prevalence of *K. pneumoniae* in different zones was significantly higher (*p*-value=0.0081).

## 4. Discussion

Our study has established a substantial presence of AMR in wastewater during the COVID-19 pandemic. Consequently, the COVID-19 pandemic and AMR are parallel and interactive health emergencies requiring further research [27]. While several clinical studies were reported from India, a very limited number were reported on environmental wastewater. Specifically, none of the studies were reported from non-metropolitan cities. Our findings revealed that almost equal MDR had prevailed during the pandemic in both metropolitan and non-metropolitan cities. Our results showed that in non-metropolitan cities too, the indiscriminate use of antibiotics to treat secondary bacterial infections and their improper disposal in the environment may have led to increased AMR concentration in the environment. This could be confirmed by our findings where an extremely significant resistance pattern for different antibiotic classes for various cities was observed in the bacterial population. As India does not have well wastewater treatment systems it would be expected that high AMR would prevail in these drainage systems that might spread to the environment and cause a risk to human health. Previously, high concentrations of biocides were used and identified in wastewater and drainage systems. Their disposal creates selective pressure on microbiota and thus increases AMR concentration in the environment, posing a human health risk for individuals exposed to these environments [40]. Through our findings we can strongly underline the role of *Enterobacteriaceae* members as vectors for the propagation of resistance and the beta-lactamases in water bodies as a high percentage of MDR was detected in *E. coli* (57.14%), followed by *K. pneumoniae* (28.57%). The use of antibiotics in India during COVID-19 has moved away from antimicrobial management, affecting AMR levels in wastewater. It is well known that untreated wastewater from hospital settings and sewage is a massive reservoir of antimicrobial-resistant strain [28,29].

Studies pointed to untreated wastewater as a possible source of MDR strains. ESBL and carbapenem-resistant mobile elements in the gut microbiota play a crucial role in MDR transfer from the environment to humans through the food chain [30]. Our findings showed 79.3% ESBL and 36.05% carbapenem-resistance in *Enterobacteriales*. *Enterobacteriaceae* harboring cefotaxime enzymes are the most prominent bacteria associated with ESBL resistance worldwide [31]. Our findings show a 74% multidrug-resistant *Enterobacteriaceae* prevalence in wastewater, supporting previous studies’ results. *E. coli* isolates obtained from hospital sewage in north-western Greece were identified as multidrug-resistant. In addition, 79.2% of *K. pneumoniae* isolates were resistant to carbapenem, and a single isolate was resistant to tigecycline [32]. A related study was conducted on wastewater in Germany in which *E. coli* contained 57% MDR, and *K. pneumonia* showed 100% MDR [33]. Furthermore, a study on hospital wastewater in New Delhi showed a high prevalence of bacterial strains resistant to ESBL and carbapenem. Approximately 18-41% of carbapenem-resistant *Enterobacteriaceae* isolates from wastewater were identified, of which 55% of isolates carried at least one *bla*_NDM-1_ gene. Hospital wastewater had a nine-fold greater carbapenem-resistant *Enterobacteriaceae* and bla_NDM-1_ gene concentration than local sewage. Thus, hospitals contribute to higher antimicrobial resistance concentrations relative to Delhi community sources, especially during winter [34]. Similarly, our findings showed carbapenem resistance (37-59%) in *Enterobacterales*. This carbapenem resistance rate when compared to previous studies denotes that during the COVID-19 pandemic the resistance rate accelerated.

It was recently observed that antimicrobial resistance in environmental water bodies in India increased in the year 2020 compared to 2018 which is in accordance with our findings. Also, antibiotic-resistant *E.* c*oli* in non-fluoroquinolone drugs is higher than in quinolones [35]. In our findings, the highest resistance was observed against penicillin, cephalosporin, quinolone, and fluoroquinolone. A study in the United States showed that 80% of MDR *Enterobacteriaceae* isolates were resistant to carbapenem, with the highest resistance to ertapenem in wastewater, similar to our findings. In our study, carbapenem resistance was higher in India compared to previous reports and is significantly higher for *K.pneumoniae* [36]. In addition, related research on 47 MDR isolates, particularly carbapenem-resistant and ESBL-producing *K*. *pneumonia* isolates, was reported from WWTPs of Romanian hospital units. Further, resistance to aminoglycoside, quinolone and sulphonamide was also observed [37]. Recent findings suggested that multidrug resistance is slightly higher in hospital wastewater than in community wastewater, meaning little significant difference. This corroborates our findings, and might have occurred because of non-regulatory improper disposal of antibiotics and biocides in the drainage system during the pandemic. As India does not have proper wastewater treatment systems, in the future this may pose a health risk to not only humans but to the whole ecosystem. The frequency of resistance varies between the two, with community isolates showing the highest resistance of *E. coli* to carbapenem (76.47% *vs* 68.09%), compared to hospital isolates where *E. coli* showed the highest resistance to ESBL (21.28% *vs* 5.88%) [38]. Exposure to antimicrobials in ecological settings is expected to exert selective pressure on the microbiota. Gene transfer through mobile elements such as integrons, plasmids, transposons, and intercellular insertion sequences, facilitates the production of extended-spectrum beta-lactamases in *Enterobacteriaceae* [39,40]. Studies reported combining resistance to ampicillin and tetracycline in ESBL-producers due to genes encoding resistance to these antibiotics located on the same plasmid [41].

Recently, a study on wastewater from India and Sri Lanka showed higher resistance to quinolones, especially ciprofloxacin, than to β-lactams and sulfonamides, which agrees with our findings [42]. Our findings reveal a combined resistance of 67.5% against ciprofloxacin for both *E. coli* and *K. pneumonia* bacterial strains. *E. coli* and *K. pneumoniae* isolated from fish in Assam (India) showed MDR against 12 antibiotics. The study emphasised aquaculture resistance that is directly or indirectly related to our findings. In *E. coli*, 6 to 100% resistance was observed against ceftazidime and cefotaxime (third-generation cephalosporin) and 85% against cefepime (fourth-generation cephalosporin). However, in our study, *E. coli* showed 66.6-100% resistance to cefotaxime compared to 83 to 100% resistance in different regions. Similarly, *K. pneumoniae* showed 100% resistance to cefotaxime, 67% resistance to ceftazidime, and 75% resistance to cefepime, which differs from our findings. We found 66.6% resistance to cefotaxime and 33-66.6% resistance to cefepime [43]. Besides these studies, many studies from India showed MDR and ESBL production in *E. coli*, *Klebsiella* and several other isolates obtained from highly polluted water sites. In one recent study, about 60% of the *Enterobacteriaceae* isolates screened were multidrug-resistant, of which 16.6% were carbapenem-resistant. All NDM-1 isolates were resistant to carbapenem, cephalosporin, aminoglycosides, quinolone, tetracycline, and sulphonamides. In contrast, our study’s resistance rate (74% MDR in *Enterobacteriaceae* isolates) was higher. This strongly suggests and links our findings of AMR with the COVID-19 pandemic. In addition, resistance to other antibiotic classes correlates [44]. Previously, a study on the prevalence of antimicrobial-resistant *E.coli* in several sewage treatment plants in South India showed antimicrobial resistance ranging from 14 to 80% against 12 antibiotics. The highest resistance rates were against ampicillin, cefotaxime, cefazolin, ciprofloxacin and nalidixic acid. In contrast, <14% resistance was recorded against imipenem and chloramphenicol. This indicates that WWTPs cannot remove MDR from wastewater [45]. A similar study showed that wastewater treatment plants could reduce *E. coli* numbers but not selectively eradicate antimicrobial-resistant strains [46]. Our findings showed that colistin resistance is emerging in other members of the *Enterobacteriaceae* family, such as *Pantoea* spp. (Table 3). This increase in resistance to colistin, the latest drug currently available to treat infections caused by MDR bacteria, is alarming. To the best of our knowledge, particularly for the above three isolates, our report is the first of its kind in the region. Therefore, our study confirmed various resistance profiles in water bodies and indicated a health risk to humans upon exposure. The indiscriminate use of antibiotics and their direct or indirect (*via* faeces) elimination in water bodies remains a cause for concern both in the past and today. Thus, the rapid increase of AMR in the environment during the COVID-19 pandemic is worrying, and strict measures must be taken. Without appropriate measures to mitigate the growing AMR threat, the COVID-19 pandemic and AMR may remain parallel and interactive health emergencies causing collateral public health damage requiring further research.

India is an emerging economy but around 60% of the sector is unorganised as per the published reports and panel discussions. Metropolitan cities are getting more advanced infrastructure facilities but the non-metropolitan cities or, so-called B class cities, which will be the future upcoming metro cities of India are far away from those important industrial infrastructure. In these non-metropolitan cities around 75% of the wastewater is disposed of without any treatment and even get released into rivers. We need to be aware that this is a problem that is not limited to a single country, but needs to be approached from a one-health perspective as a global problem, as resistance moves rapidly from country to country [53]. With our research findings, at least we can draw the attention of our current government, policy makers, the scientific community and funding scientific agencies to rethink the problem and draft a more suitable policy before upgrading the infrastructure facilities in other Indian cities.

## 5. Conclusion

The present study reveals the presence of *Enterobacteriaceae* isolates with diverse antibiotic resistance patterns in different wastewater samples in India, including resistance to last-resort antibiotics like colistin. Approximately 80% of the isolates were multidrug-resistant. These findings suggest that antibiotic use in India during COVID-19 has moved away from antimicrobial stewardship, and the level of AMR in wastewater is bound to increase soon. Considering that India is a growing country with about one-fifth of the world’s entire population, it is easy to see that the possibility of bacterial resistance transferring to other countries is increasing and that it is increasingly a global problem. A more in-depth work needs to be done at ground level. Health issues are arising at higher level in almost all metropolitan and non-metropolitan cities in India, however, the data is not disclosed properly especially for non-metropolitan cities. Therefore, it is suggested that periodic surveys be carried out to study the patterns and prevalence of AMR in Indian wastewater and their receiving water bodies. Furthermore, studies on the molecular characterization of antibiotic resistance genes in these areas should be conducted. Such information would improve the understanding of epidemiology and the spread of AMR in urban and rural areas of India.

## Supplementary Materials

Table S1: Biochemical characterization of *Enterobacteriaceae* isolates obtained from wastewater of Western Uttar Pradesh

Table S2: Antimicrobial susceptibility testing VITEK results.

## Author Contributions

Conceptualization, H.Q. and T.H.; Methodology, H.Q. and T.H.; Software, H.Q. and T.H.; Validation, H.Q. and T.H.; Investigation, H.Q. and T.H.; Resources, H.Q. and S.B.J; Data curation, H.Q., T.H.; Writing—original draft preparation, H.Q.; Writing—review and editing, H.Q., T.H.; Visualization, H.Q., T.H.; Supervision, S.B.J.; Project administration, H.Q.; Funding acquisition, H.Q.; All authors have read and agreed to the published version of the manuscript.

## Institutional Review Board Statement

Not applicable

## Data Availability

All data produced in the present study are available upon reasonable request to the authors

## Acknowledgments

The authors are highly thankful to the Department of Science and Technology (DST), New Delhi, India, for sanctioning the project. Thanks also to Dr. Midhat, Ms. Nikhat, Ms. Anam and all my lab mates for helping me throughout the study. The authors are highly thankful to IBU and JNMC of AMU, Aligarh, for providing infrastructure and other facilities for conducting this project. Finally, the first author is highly grateful to Dr.Nameeta Gupta (Advisor/Scientist G at DST), Dr.Chhama Awasthi, Dr.Pawan Kumar (Scientist C, KIRAN Division, DST), and all the expert committee members of DST for giving me the opportunity to initiate this work and providing lab space and a mentor at IBU, AMU, Aligarh.

## References

1. Pourajam, S.; Kalantari, E.; Talebzadeh, H.; Mellali, H.; Sami, R.; Soltaninejad, F.; Amra, B.; Sajadi, M.; Alenaseri, M.; Kalantari, F.; Solgi, H. Secondary Bacterial Infection and Clinical Characteristics in Patients With COVID-19 Admitted to Two Intensive Care Units of an Academic Hospital in Iran During the First Wave of the Pandemic. 2022. Front. Cell. Infect. Microbiol., 12.

2. De Bruyn, A.; Verellen, S.; Bruckers, L.; Geebelen, L.; Callebaut, I.; De Pauw, I.; Stessel, B.; Dubois, J. 2022. Secondary infection in COVID-19 critically ill patients: a retrospective singlecenter evaluation. BMC Infect. Dis., 22, 1–7.

3. Rawson, T. M.; Ming, D.; Ahmad, R.; Moore, L. S. P.; Holmes, A. H. 2020. Antimicrobial use, drug-resistant infections and COVID-19. Nat. Rev. Microbiol., 18, 409–410.

4. Pulingam, T.; Parumasivam, T.; Gazzali, A. M.; Sulaiman, A. M.; Chee, J. Y.; Lakshmanan, M.; Chin, C. F.; Sudesh, K. 2022. Antimicrobial resistance: Prevalence, economic burden, mechanisms of resistance and strategies to overcome. Eur. J. Pharm. Sci., 170, 106103.

5. Ahmad, M.; Khan, A. U. 2019. Global economic impact of antibiotic resistance: A review. J. Glob. Antimicrob. Resist., 19, 313–316.

6. Manesh, A.; Varghese, G. M. 2021. Rising antimicrobial resistance: an evolving epidemic in a pandemic. The Lancet Microbe, 2, e419–e420.

7. Sulis, G.; Batomen, B.; Kotwani, A.; Pai, M.; Gandra, S. 2021. Sales of antibiotics and hydroxychloroquine in India during the COVID-19 epidemic: An interrupted time series analysis. PLOS Med., 18, e1003682.

8. Sulis, G.; Batomen, B.; Kotwani, A.; Pai, M.; Gandra, S. 2021. Sales of antibiotics and hydroxychloroquine in India during the COVID-19 epidemic: An interrupted time series analysis. PLOS Med., 18, e1003682.

9. Russell, C. D.; Fairfield, C. J.; Drake, T. M.; Turtle, L.; Seaton, R. A.; Wootton, D. G.; Sigfrid, L.; Harrison, E. M.; Docherty, A. B.; de Silva, T. I.; et al. 2021. Co-infections, secondary infections, and antimicrobial use in patients hospitalised with COVID-19 during the first pandemic wave from the ISARIC WHO CCP-UK study: a multicentre, prospective cohort study. The Lancet Microbe, 2, e354–e365.

10. WHO. 2017. *Global priority list of antibiotic-resistant batceria to guide research, discovery, and development of new antibiotics*; Geneva, Switzerland.

11. Lynch, J. P.; Clark, N. M.; Zhanel, G. G. 2021. Escalating antimicrobial resistance among Enterobacteriaceae: focus on carbapenemases. Expert Opin. Pharmacother., 22, 1455–1473.

12. Falagas, M. E.; Karageorgopoulos, D. E. 2009. Extended-spectrum β-lactamase-producing organisms. J. Hosp. Infect, 73, 345–354.

13. Kagambèga, A.; Haukka, K.; Siitonen, A.; Traoré, A. S.; Barro, N. 2011. Prevalence of Salmonella enterica and the hygienic indicator Escherichia coli in raw meat at markets in Ouagadougou, Burkina Faso. J. Food Prot., 74, 1547–1551.

14. Jakobsen, L.; Kurbasic, A.; Skjøt-Rasmussen, L.; Ejrnæs, K.; Porsbo, L. J.; Pedersen, K.; Jensen, L. B.; Emborg, H.-D.; Agersø, Y.; Olsen, K. E. P. 2010. Escherichia coli isolates from broiler chicken meat, broiler chickens, pork, and pigs share phylogroups and antimicrobial resistance with community-dwelling humans and patients with urinary tract infection. Foodborne Pathog. Dis., 7, 537–547.

15. Yang, S. C.; Lin, C. H.; Aljuffali, I. A.; Fang, J. Y. 2017. Current pathogenic Escherichia coli foodborne outbreak cases and therapy development. Arch. Microbiol., 199, 811–825.

16. Liu, C. M.; Stegger, M.; Aziz, M.; Johnson, T. J.; Waits, K.; Nordstrom, L.; Gauld, L.; Weaver, B.; Rolland, D.; Statham, S.; Horwinski, J.; Sariya, S.; Davis, G. S.; Sokurenko, E.; Keim, P.; Johnson, J. R.; Price, L. B. 2018. Escherichia coli ST131-H22 as a Foodborne Uropathogen. MBio, 9.

17. Yang, X.; Sun, Q.; Li, J.; Jiang, Y.; Li, Y.; Lin, J.; Chen, K.; Chan, E. W. C.; Zhang, R.; Chen, S. 2022. Molecular epidemiology of carbapenem-resistant hypervirulent Klebsiella pneumoniae in China. Emerg. Microbes Infect., 11, 841–849.

18. Schwarz, S.; Johnson, A. P. 2016. Transferable resistance to colistin: A new but old threat. J. Antimicrob. Chemother., 71, 2066–2070.

19. Nguyen, N. T.; Nguyen, H. M.; Nguyen, C. V; Nguyen, T. V; Nguyen, M. T.; Thai, H. Q.; Ho, M. H.; Thwaites, G.; Ngo, H. T.; Baker, S.; Carrique-Mas, J. 2016. Use of Colistin and Other Critical Antimicrobials on Pig and Chicken Farms in Southern Vietnam and Its Association with Resistance in Commensal Escherichia coli Bacteria. Appl Env. Microbiol, 82, 3727–3735.

20. Liu, Y. Y.; Wang, Y.; Walsh, T. R.; Yi, L. X.; Zhang, R.; Spencer, J.; Doi, Y.; Tian, G.; Dong, B.; Huang, X.; Yu, L. F.; Gu, D.; Ren, H.; Chen, X.; Lv, L.; He, D.; Zhou, H.; Liang, Z.; Liu, J. H.; Shen, J. 2016. Emergence of plasmid-mediated colistin resistance mechanism MCR-1 in animals and human beings in China: a microbiological and molecular biological study. Lancet Infect. Dis., 16, 161–168.

21. Sun, J.; Zhang, H.; Liu, Y.-H.; Feng, Y. 2018. Towards understanding MCR-like colistin resistance. Trends Microbiol., 26, 794–808.

22. Shein, A. M. S.; Hongsing, P.; Abe, S.; Luk-in, S.; Ragupathi, N. K. D.; Wannigama, D. L.; Chatsuwan, T. 2021. Will There Ever Be Cure for Chronic, Life-Changing Colistin-Resistant Klebsiella pneumoniae in Urinary Tract Infection? Front. Med., 8.

23. Bengtsson-Palme, J.; Hammarén, R.; Pal, C.; Östman, M.; Björlenius, B.; Flach, C. F.; Fick, J.; Kristiansson, E.; Tysklind, M.; Larsson, D. G. J. 2016. Elucidating selection processes for antibiotic resistance in sewage treatment plants using metagenomics. Sci. Total Environ, 572, 697–712.

24. Batt, A. L.; Bruce, I. B.; Aga, D. S. 2006. Evaluating the vulnerability of surface waters to antibiotic contamination from varying wastewater treatment plant discharges. Environ. Pollut.142, 295–302.

25. Sinthuchai, D.; Boontanon, S. K.; Boontanon, N.; Polprasert, C. 2016.Evaluation of removal efficiency of human antibiotics in wastewater treatment plants in Bangkok, Thailand. Water Sci. Technol., 73, 182–91.

26. Schoeman, C.; Dlamini, M.; Okonkwo, O. J. 2017. The impact of a Wastewater Treatment Works in Southern Gauteng, South Africa on efavirenz and nevirapine discharges into the aquatic environment. Emerg. Contam., 3, 95–106.

27. Ukuhor, H. O. 2021. The interrelationships between antimicrobial resistance, COVID-19, past, and future pandemics. J. Infect. Public Health, 14, 53–60.

28. Ogura, Y.; Ueda, T.; Nukazawa, K.; Hiroki, H.; Xie, H.; Arimizu, Y.; Hayashi, T.; Suzuki, Y. 2020. The level of antimicrobial resistance of sewage isolates is higher than that of river isolates in different Escherichia coli lineages. Sci. Rep., 10, 1–10.

29. Hunter, P. R.; Wilkinson, D. C.; Catling, L. A.; Barker, G. C. 2008. Meta-analysis of experimental data concerning antimicrobial resistance gene transfer rates during conjugation. Appl. Environ. Microbiol, 74, 6085–6090.

30. Blaak, H.; De Kruijf, P.; Hamidjaja, R. A.; Van Hoek, A. H. A. M.; De Roda Husman, A. M.; Schets, F. M. 2014. Prevalence and characteristics of ESBL-producing E. coli in Dutch recreational waters influenced by wastewater treatment plants. Vet. Microbiol., 171, 448–459.

31. Franz, E.; Veenman, C.; Van Hoek, A. H. A. M.; Husman, A. D. R.; Blaak, H. 2015. Pathogenic Escherichia coli producing Extended-Spectrum β-Lactamases isolated from surface water and wastewater. Sci. Rep., 5, 14372.

32. Sakkas, H.; Bozidis, P.; Ilia, A.; Mpekoulis, G.; Papadopoulou, C. 2019. Antimicrobial resistance in bacterial pathogens and detection of carbapenemases in klebsiella pneumoniae isolates from hospital wastewater. Antibiotics, 8.

33. Homeier-bachmann, T.; Heiden, S. E.; Lübcke, P. K.; Bachmann, L.; Bohnert, J. A.; Zimmermann, D.; Schaufler, K. 2021. Antibiotic-Resistant Enterobacteriaceae in Wastewater of Abattoirs.

34. Lamba, M.; Graham, D. W.; Ahammad, S. Z. 2017. Hospital Wastewater Releases of Carbapenem-Resistance Pathogens and Genes in Urban India. Environ. Sci. Technol., 51, 13906–13912.

35. Kumar, M.; Dhangar, K.; Thakur, A. K.; Ram, B.; Chaminda, T.; Sharma, P.; Kumar, A.; Raval, N.; Srivastava, V.; Rinklebe, J.; Kuroda, K.; Sonne, C.; Barcelo, D. 2021. Antidrug Resistance in the Indian Ambient Waters of Ahmedabad during the COVID-19 Pandemic. J. Hazard. Mater., 416, 126125.

36. Reinke, R. A.; Quach-Cu, J.; Allison, N.; Lynch, B.; Crisostomo, C.; Padilla, M. 2020. A method to quantify viable carbapenem resistant gram-negative bacteria in treated and untreated wastewater. J. Microbiol. Methods, 179, 106070.

37. Surleac, M.; Barbu, I. C.; Paraschiv, S.; Popa, L. I.; Gheorghe, I.; Marutescu, L.; Popa, M.; Sarbu, I.; Talapan, D.; Nita, M.; Iancu, A. V.; Arbune, M.; Manole, A.; Nicolescu, S.; Sandulescu, O.; Streinu-Cercel, A.; Otelea, D.; Chifiriuc, M. C. 2020. Whole genome sequencing snapshot of multidrug resistant Klebsiella pneumoniae strains from hospitals and receiving wastewater treatment plants in Southern Romania. PLoS One, 15, 1–17.

38. Gaşpar, C. M.; Cziszter, L. T.; Lăzărescu, C. F.; Ţibru, I.; Pentea, M.; Butnariu, M. 2021. Antibiotic resistance among escherichia coli isolates from hospital wastewater compared to community wastewater. Water (Switzerland), 13, 1–11.

39. Subramanya, S. H.; Bairy, I.; Metok, Y.; Baral, B. P.; Gautam, D.; Nayak, N. 2021. Detection and characterization of ESBL-producing Enterobacteriaceae from the gut of subsistence farmers, their livestock, and the surrounding environment in rural Nepal. Sci. Rep, 11, 1–13.

40. Murray, B. E. 1992. Problems and Dilemmas of Antimicrobial Resistance. Pharmacotherapy, 12, 86S–93S.

41. Nys, S.; Okeke, I. N.; Kariuki, S.; Dinant, G. J.; Driessen, C.; Stobberingh, E. E. 2004. Antibiotic resistance of faecal Escherichia coli from healthy volunteers from eight developing countries. J. Antimicrob. Chemother., 54, 952–955.

42. Kumar, M.; Ram, B.; Sewwandi, H.; Sulfikar Honda, R.; Chaminda, T. 2020. Treatment enhances the prevalence of antibiotic-resistant bacteria and antibiotic resistance genes in the wastewater of Sri Lanka, and India. Environ. Res., 183.

43. Sivaraman, G. K.; Sudha, S.; Muneeb, K. H.; Shome, B.; Holmes, M.; Cole, J. 2020. Molecular assessment of antimicrobial resistance and virulence in multi drug resistant ESBL-producing Escherichia coli and Klebsiella pneumoniae from food fishes, Assam, India. Microb. Pathog., 149, 104581.

44. Kalasseril, S. G.; Krishnan, R.; Vattiringal, R. K.; Paul, R.; Mathew, P.; Pillai, D. 2020. Detection of New Delhi Metallo-β-lactamase 1 and Cephalosporin Resistance Genes Among Carbapenem-Resistant Enterobacteriaceae in Water Bodies Adjacent to Hospitals in India. Curr. Microbiol., 77, 2886–2895.

45. Akiba, M.; Senba, H.; Otagiri, H.; Prabhasankar, V. P.; Taniyasu, S.; Yamashita, N.; Lee, Kichi; Yamamoto, T.; Tsutsui, T.; Ian Joshua, D.; Balakrishna, K.; Bairy, I.; Iwata, T.; Kusumoto, M.; Kannan, K.; Guruge, K. S. 2015. Impact of wastewater from different sources on the prevalence of antimicrobial-resistant Escherichia coli in sewage treatment plants in South India. Ecotoxicol. Environ. Saf., 115, 203–208.

46. Korzeniewska, E.; Korzeniewska, A.; Harnisz, M. 2013. Antibiotic resistant Escherichia coli in hospital and municipal sewage and their emission to the environment. Ecotoxicol. Environ. Saf., 91, 96–102.

47. Hassen, B.; Abbassi, M. S.; Benlabidi, S.; Ruiz-Ripa, L.; Mama, O. M.; Ibrahim, C.; Hassen, A.; Hammami, S.; Torres, C. 2020. Genetic characterization of ESBL-producing Escherichia coli and Klebsiella pneumoniae isolated from wastewater and river water in Tunisia: predominance of CTX-M-15 and high genetic diversity. Environ. Sci. Pollut. Res., 27, 44368–44377.

48. Tiwari, R.; Nayak, S. 2013. Drinking water and sanitation in Uttar Pradesh: A regional analysis. J. Rural Dev., 32, 61–74.

49. Bauer, A.; Kirby, W.; Sherris, J.; Turck, M. 1966. Antibiotic susceptibility testing by a standardised single disk method. Am. J. Clin. Pathol., 45, 413–496.

50. CLSI CLSI M100-ED30:2020 Performance Standards for Antimicrobial Susceptibility Testing, 30th Edition; Wayne, PA, 2020.

51. Simner, P. J.; Bergman, Y.; Trejo, M.; Roberts, A. A.; Marayan, R.; Tekle, T.; Campeau, S.; Kazmi, A. Q.; Bell, D. T.; Lewis, S.; Tamma, P. D.; Humphries, R. M.; Hindler, J. A. 2019. Two-Site Evaluation of the Colistin Broth Disk Elution Test To Bacilli. J. Clin. Microbiol., 57, 1–7.

52. Murray, A.K.;2020. The Novel Coronavirus COVID-19 Outbreak: Global Implications for Antimicrobial Resistance. Front. Microbiol., 11:1020. doi: 10.3389/fmicb.2020.01020

53. Struelens MJ, Monnet DL, Magiorakos AP, Santos O’Connor F, Giesecke J, the European NDM-1 Survey Participants. New Delhi metallo-beta-lactamase 1–producing Enterobacteriaceae: emergence and response in Europe. Euro Surveill. 2010;15(46):pii=19716. Available online: http://www.eurosurveillance.org/ViewArticle.aspx?ArticleId=19716

