## supplementary file 1 for "Urban Wastewater as Environmental Reservoirs of Multidrug-resistance *Enterobacteriaceae* during the COVID-19 Pandemic in India"

**Supplementary Data:**

**Table 1(a): Biochemical characterization of Enterobacteriaceae isolates obtained from waste water of western U.P.**

| Number of Isolates (n=150) | ONPG | Lysine | Ornithine | Urease | TDA | Nitrate | $H_2S$ | Citrate Utilization | Voge's Proskauer's | Methyl Red | Indole | Malonate | Result Interpretations |
|---|---|---|---|---|---|---|---|---|---|---|---|---|---|
| | | | | | | ZONE 1 | | | | | | | |
| 1G | ++ | ++ | + | - | - | ++ | - | - | - | ++ | ++ | - | *E. coli* |
| 2 G | ++ | ++ | - | ++ | - | + | - | ++ | + | + | - | ++ | *K. pneumonia* |
| 3 G | ++ | ++ | - | ++ | - | ++ | - | ++ | ++ | + | - | ++ | *K. pneumoniae* |
| 4G | - | ++ | + | - | - | + | - | - | - | ++ | ++ | - | *E. coli*(S) |
| 5G | + | + | + | - | - | ++ | - | - | - | ++ | + | - | *E. coli*(S) |
| 6G | + | + | + | - | - | ++ | - | - | - | ++ | + | - | *E. coli*(S) |
| 7G | ++ | ++ | - | ++ | - | ++ | - | ++ | ++ | + | - | ++ | *K. pneumoniae*(C) |
| 8G | - | ++ | + | - | - | + | - | - | - | ++ | + | - | *E. coli*(S) |
| 9G | - | - | + | - | + | + | - | - | ++ | + | + | - | Mixed population |
| 10G | + | + | + | - | - | ++ | - | - | - | ++ | + | - | *E. coli*(S) |
| 11G | - | - | - | - | - | ++ | - | - | - | ++ | + | - | *Shigella* (S) |
| 12G | ++ | ++ | + | - | - | ++ | - | - | - | ++ | ++ | - | *E. coli*(C) |
| 13G | + | + | + | - | - | + | - | - | - | ++ | + | - | *E. coli* (S) |
| 14G | ++ | ++ | - | ++ | - | ++ | - | ++ | ++ | + | - | ++ | *K. pneumoniae*(S) |
| 15G | + | + | + | - | - | ++ | - | - | - | ++ | + | - | *E. coli*(S) |
| 16G | - | ++ | + | - | - | + | - | - | - | ++ | + | - | *E. coli*(S) |
| 17G | ++ | ++ | - | ++ | - | ++ | - | ++ | ++ | + | - | + | *K. pneumoniae*(S) |
| 18G | - | ++ | + | - | - | + | - | - | - | + | + | - | *E. coli*(S) |
| 19G | ++ | + | - | ++ | - | ++ | - | ++ | ++ | + | - | ++ | *K. pneumoniae*(S) |
| 20G | ++ | ++ | + | - | - | ++ | - | - | - | ++ | ++ | - | *E. coli*(C) |
| 21N | - | - | - | - | - | ++ | - | - | - | ++ | + | - | *Shigella* (S) |
| 22N | ++ | ++ | - | ++ | - | ++ | - | ++ | ++ | + | - | ++ | *K. pneumoniae*(S) |
| 23N | ++ | ++ | + | - | - | ++ | - | - | - | ++ | ++ | - | *E. coli*(S) |
| 24N | + | + | + | - | - | ++ | - | - | - | ++ | + | - | *E. coli*(S) |
| 25N | ++ | ++ | - | ++ | - | ++ | - | ++ | + | + | - | ++ | *K. pneumoniae*(S) |
| 26N | - | ++ | + | - | - | + | - | - | - | ++ | ++ | - | *E. coli*(S) |
| 27N | - | - | ++ | - | * | - | - | ++ | * | * | - | - | *Alcaligenes faecalis*(S) |
| 28N | + | + | + | - | - | ++ | - | - | - | ++ | + | - | *E. coli*(S) |

| | | | | | | | | | | | | | |
|---|---|---|---|---|---|---|---|---|---|---|---|---|---|
| 29N | ++ | ++ | + | - | - | ++ | - | - | - | ++ | ++ | - | *E. coli*(C) |
| 30N | - | + | + | - | - | ++ | - | - | - | + | + | - | *E. coli*(S) |
| 31D | ++ | ++ | - | ++ | - | ++ | - | ++ | ++ | + | - | ++ | *K. pneumonia* |
| 32D | - | ++ | + | - | - | + | - | - | - | + | + | - | *E. coli*(S) |
| 33D | - | + | - | - | - | + | - | - | - | + | + | - | *E. coli*(S) |
| 34D | ++ | ++ | - | ++ | - | ++ | - | ++ | ++ | + | - | ++ | *K. pneumonia*(C) |
| 35D | ++ | ++ | - | ++ | - | ++ | - | ++ | ++ | + | - | + | *K. pneumonia*(S) |
| 36D | - | ++ | + | - | - | + | - | - | - | + | + | - | *E. coli*(S) |
| 37D | - | + | ++ | - | - | - | - | - | + | + | + | + | Mixed population |
| 38D | + | + | + | - | - | ++ | - | - | - | ++ | + | - | *E. coli*(S) |
| 39D | ++ | ++ | - | ++ | - | ++ | - | ++ | ++ | + | - | ++ | *K. pneumonia*(S) |
| 40D | - | - | ++ | - | * | - | - | ++ | * | * | - | - | *Alcaligenes faecalis*(S) |
| 41D | ++ | ++ | + | - | - | ++ | - | - | - | ++ | ++ | - | *E. coli*(C) |
| 42D | + | + | + | - | - | ++ | - | - | - | ++ | + | - | *E. coli*(S) |
| 43D | + | + | + | - | - | ++ | - | - | - | ++ | + | - | *E. coli*(S) |
| 44D | ++ | ++ | + | - | - | ++ | - | - | - | ++ | ++ | - | *E. coli*(S) |
| 45D | ++ | ++ | + | - | - | ++ | - | - | - | ++ | ++ | - | *E. coli*(C) |
| 46D | ++ | ++ | - | ++ | - | ++ | - | ++ | ++ | + | - | ++ | *K. pneumoniae*(S) |
| 47D | - | ++ | + | - | - | + | - | - | - | + | + | - | *E. coli*(S) |
| 48D | ++ | + | - | ++ | - | ++ | - | ++ | ++ | + | - | ++ | *K. pneumoniae*(S) |
| 49D | - | ++ | + | - | - | + | - | - | - | + | + | - | *E. coli*(S) |
| 50D | - | - | ++ | - | * | - | - | ++ | * | * | - | - | *Alcaligenes faecalis*(S) |
| **ZONE 2** | | | | | | | | | | | | | |
| 51A | ++ | ++ | + | - | - | ++ | - | - | - | ++ | ++ | - | *E. coli*(S) |
| 52A | ++ | ++ | + | - | - | ++ | - | - | - | ++ | ++ | - | *E. coli*(C) |
| 53A | + | + | + | - | - | ++ | - | - | - | ++ | + | - | *E. coli*(S) |
| 54A | ++ | ++ | - | ++ | - | ++ | - | ++ | ++ | + | - | ++ | *K. pneumonia*(C) |
| 55A | ++ | ++ | - | ++ | - | ++ | - | ++ | ++ | + | - | ++ | *K. pneumoniae*(C) |
| 56A | + | ++ | + | - | - | + | - | - | - | + | + | - | *E. coli*(S) |
| 57A | + | + | + | + | - | + | - | - | + | + | + | + | Mixed population |
| 58A | ++ | ++ | + | - | - | ++ | - | - | - | ++ | ++ | - | *E. coli*(C) |
| 59A | - | ++ | + | - | - | ++ | - | - | - | + | + | - | *E. coli*(S) |
| 60A | - | ++ | + | - | - | + | - | - | - | ++ | + | - | *E. coli*(S) |
| 61A | - | - | ++ | - | * | - | - | ++ | * | * | - | - | *Alcaligenes faecalis*(S) |

| | | | | | | | | | | | | | |
|---|---|---|---|---|---|---|---|---|---|---|---|---|---|
| 62A | - | - | - | ++ | - | ++ | - | ++ | ++ | + | + | + | *K. pneumoniae*(S) |
| 63A | + | + | + | - | - | ++ | - | - | - | ++ | + | - | *E. coli*(S) |
| 64A | + | + | + | - | - | ++ | - | - | - | ++ | + | - | *E. coli*(S) |
| 65A | + | - | - | ++ | - | ++ | - | ++ | ++ | + | - | + | *K. pneumoniae*(S) |
| 66A | + | ++ | + | - | - | + | - | - | + | + | + | + | Mixed population |
| 67A | ++ | ++ | + | - | - | + | - | - | - | + | + | - | *E. coli*(S) |
| 68A | + | + | + | - | - | ++ | - | - | - | ++ | + | - | *E. coli*(S) |
| 69A | - | - | ++ | - | * | - | - | ++ | * | * | - | - | *Alcaligenes faecalis*(S) |
| 70A | + | ++ | + | - | - | + | - | - | - | ++ | + | - | *E. coli*(S) |
| 71Al | + | + | + | - | - | + | - | - | - | + | + | - | *E. coli*(S) |
| 72Al | ++ | ++ | - | ++ | - | ++ | - | ++ | ++ | + | - | ++ | *K. pneumoniae*(C) |
| 73Al | ++ | ++ | + | - | - | ++ | - | - | - | ++ | ++ | - | *E. coli*(S) |
| 74Al | ++ | ++ | + | - | - | ++ | - | - | - | ++ | ++ | - | *E. coli*(C) |
| 75Al | + | ++ | + | - | - | + | - | - | - | + | + | - | *E. coli*(S) |
| 76Al | - | - | - | ++ | - | ++ | - | ++ | ++ | + | - | ++ | *K. pneumonia* (S) |
| 77Al | ++ | - | + | - | + | ++ | - | ++ | ++ | + | - | ++ | *Pantoea agglomerans* (C) |
| 78Al | + | ++ | + | - | - | + | - | - | - | + | + | - | *E. coli*(S) |
| 79Al | ++ | ++ | + | - | - | ++ | - | - | - | ++ | ++ | - | *E. coli*(C) |
| 80Al | - | - | ++ | - | * | - | - | ++ | * | * | - | - | *Alcaligenes faecalis*(S) |
| 81Al | - | ++ | + | - | - | + | - | - | - | + | + | - | *E. coli*(S) |
| 82Al | - | ++ | + | - | - | ++ | - | - | - | + | + | - | *E. coli*(S) |
| 83Al | ++ | ++ | - | ++ | - | ++ | - | ++ | ++ | + | - | ++ | *K. pneumoniae*(C) |
| 84Al | + | + | + | - | - | + | - | - | - | ++ | + | - | *E. coli*(S) |
| 85Al | - | + | ++ | - | - | ++ | - | - | - | ++ | + | - | *E. coli*(S) |
| 86Al | + | ++ | + | - | - | + | - | - | - | + | + | - | *E. coli*(S) |
| 87Al | - | - | ++ | - | * | - | - | ++ | * | * | - | - | *Alcaligenes faecalis*(S) |
| 88Al | - | ++ | + | - | - | + | - | - | - | + | + | - | *E. coli*(S) |
| 89Al | + | + | + | - | - | ++ | - | - | - | ++ | + | - | *E. coli*(S) |
| 90Al | + | + | + | - | - | ++ | - | - | - | ++ | + | - | Mixed population |
| 91H | - | - | - | - | - | ++ | - | + | + | + | - | ++ | *K. pneumoniae*(S) |
| 92H | ++ | ++ | + | - | - | ++ | - | - | - | ++ | ++ | - | *E. coli*(C) |
| 93H | ++ | ++ | + | - | - | ++ | - | - | - | ++ | ++ | - | *E. coli*(S) |
| 94H | - | ++ | + | - | - | - | + | ++ | - | - | + | ++ | *E. coli*(S) |

| | | | | | | | | | | | | | |
|---|---|---|---|---|---|---|---|---|---|---|---|---|---|
| 95H | - | ++ | ++ | - | - | - | ++ | ++ | - | - | - | ++ | *E. coli*(S) |
| 96H | - | ++ | + | - | - | + | - | - | - | ++ | ++ | - | *E. coli*(S) |
| 97H | - | - | - | - | - | ++ | - | + | + | + | - | ++ | *K. pneumonia*(S) |
| 98H | - | ++ | + | - | - | + | - | - | - | ++ | ++ | - | *E. coli*(S) |
| 99H | - | ++ | + | + | - | + | - | ++ | - | ++ | - | - | Mixed population |
| 100H | + | + | + | - | - | ++ | - | - | - | ++ | + | - | *E. coli*(S) |
| **ZONE 3** | | | | | | | | | | | | | |
| 101M | ++ | ++ | + | - | - | ++ | - | - | - | ++ | ++ | - | *E. coli*(C) |
| 102M | ++ | ++ | + | - | - | ++ | - | - | - | ++ | ++ | - | *E. coli*(C) |
| 103M | + | + | + | - | - | + | - | + | + | + | - | ++ | *K. pneumonia*(S) |
| 104M | ++ | ++ | - | - | - | ++ | - | + | + | + | - | + | *K. pneumoniae*(S) |
| 105M | ++ | - | + | - | + | ++ | - | ++ | ++ | + | - | ++ | *Pantoea agglomerans*(S) |
| 106M | - | ++ | + | - | - | ++ | - | - | - | + | + | - | *E. coli*(S) |
| 107M | - | ++ | + | - | - | + | - | - | - | ++ | + | - | *E. coli*(S) |
| 108M | ++ | ++ | - | ++ | - | ++ | - | ++ | ++ | + | - | ++ | *K. pneumoniae*(C) |
| 109M | + | + | + | - | - | ++ | - | - | - | ++ | + | - | *E. coli*(S) |
| 110M | + | + | + | - | - | ++ | - | - | - | ++ | + | - | *E. coli*(S) |
| 111M | - | - | - | - | - | ++ | - | + | + | + | + | + | *K. pneumoniae*(S) |
| 112M | + | + | + | - | - | ++ | - | - | - | ++ | + | - | *E. coli*(S) |
| 113M | + | + | + | - | - | + | - | - | - | ++ | + | - | *E. coli*(S) |
| 114M | + | + | + | - | - | ++ | - | - | - | + | + | - | *E. coli*(S) |
| 115M | ++ | ++ | + | - | - | ++ | - | - | - | ++ | ++ | - | *E. coli*(C) |
| 116M | + | ++ | + | - | - | ++ | - | - | - | ++ | + | - | *E. coli*(S) |
| 117M | ++ | + | + | - | - | + | - | - | - | + | ++ | - | *E. coli*(S) |
| 118M | + | + | + | - | - | ++ | - | - | - | ++ | + | - | Mixed population |
| 119M | ++ | + | + | - | - | + | - | - | - | ++ | + | - | *E. coli*(S) |
| 120M | + | + | + | - | - | ++ | - | - | - | ++ | + | - | *E. coli*(S) |
| 121B | ++ | ++ | - | ++ | - | ++ | - | ++ | ++ | + | - | ++ | *K. pneumoniae*(C) |
| 122B | + | + | + | - | - | + | - | - | - | ++ | + | - | *E. coli*(S) |
| 123B | + | + | + | - | - | + | - | - | - | + | + | - | *E. coli*(S) |
| 124B | + | + | + | - | - | ++ | - | - | - | ++ | + | - | *E. coli*(S) |
| 125B | + | + | + | - | - | ++ | - | - | - | ++ | + | - | *E. coli*(S) |
| 126B | + | + | + | - | - | ++ | - | - | - | ++ | + | - | *E. coli*(S) |
| 127B | - | - | - | - | - | ++ | - | + | + | + | - | ++ | *K. pneumoniae*(S) |
| 128B | ++ | ++ | - | - | - | ++ | - | + | + | + | - | ++ | *K. pneumoniae*(S) |
| 129B | + | ++ | + | - | - | ++ | - | - | - | ++ | + | + | Mixed population |

| | | | | | | | | | | | | |
|---|---|---|---|---|---|---|---|---|---|---|---|---|
| 130B | + | + | - | - | - | + | - | + | - | + | - | - | *K. pneumoniae*(S) |
| 131B | + | + | + | - | - | ++ | - | - | - | ++ | + | - | *E. coli*(S) |
| 132B | ++ | + | - | - | - | + | - | + | - | + | - | ++ | *K. pneumoniae*(S) |
| 133B | ++ | ++ | + | - | - | ++ | - | - | - | ++ | ++ | - | *E. coli*(C) |
| 134B | ++ | ++ | + | - | - | ++ | - | - | - | ++ | ++ | - | *E. coli*(C) |
| 135B | ++ | ++ | + | - | - | ++ | - | - | - | ++ | + | - | *E. coli*(S) |
| 136B | ++ | ++ | - | ++ | - | ++ | - | ++ | ++ | + | - | ++ | *K. pneumoniae*(C) |
| 137B | + | ++ | + | - | - | ++ | - | - | - | ++ | + | - | *E. coli*(S) |
| 138B | + | + | - | - | - | + | - | + | + | + | - | ++ | *K. pneumoniae*(S) |
| 139B | ++ | ++ | + | - | - | ++ | - | - | - | ++ | ++ | - | *E. coli*(C) |
| 140B | + | + | + | - | - | + | - | - | - | + | + | - | *E. coli*(S) |
| 141Bj | + | + | - | - | - | + | - | + | - | + | - | - | *K. pneumoniae*(S) |
| 142Bj | | | | | | | | | | | | | *S. enteritidis* (S) |
| 143Bj | + | + | + | - | - | ++ | - | - | - | ++ | + | - | *E. coli*(S) |
| 144Bj | ++ | + | + | - | - | ++ | - | - | - | ++ | + | - | *E. coli*(S) |
| 145Bj | ++ | ++ | - | + | - | + | - | + | + | + | - | + | *K. pneumoniae*(S) |
| 146Bj | + | + | + | - | - | + | - | - | - | ++ | + | - | *E. coli*(S) |
| 147Bj | - | ++ | + | + | - | + | - | - | - | ++ | - | ++ | Mixed population |
| 148Bj | + | + | + | - | - | ++ | - | - | - | ++ | + | - | *E. coli*(S) |
| 149Bj | ++ | + | + | - | - | + | - | - | - | ++ | + | - | *E. coli*(S) |
| 150Bj | ++ | ++ | + | - | - | ++ | - | - | - | ++ | ++ | - | *E. coli*(C) |

| **Number of Isolates (n=150)** | **Esculin Hydrolysis** | **Arabinose** | **Xylose** | **Adonitol** | **Rhamnose** | **Cellobiose** | **Melibiose** | **Saccharose** | **Raffinose** | **Trehalose** | **Glucose** | **Lactose** | **Results Interpretations** |
|---|---|---|---|---|---|---|---|---|---|---|---|---|---|
| | | | | | | **ZONE 1** | | | | | | | |
| 1G | + | ++ | ++ | - | + | - | + | + | + | ++ | ++ | ++ | *E. coli* (C) |
| 2 G | ++ | + | ++ | ++ | + | ++ | ++ | ++ | ++ | ++ | ++ | ++ | *K. pneumonia*(S) |
| 3 G | ++ | ++ | ++ | ++ | ++ | ++ | ++ | ++ | ++ | ++ | ++ | ++ | *K. pneumoniae*(C) |
| 4G | - | + | + | - | + | - | + | + | + | ++ | ++ | + | *E. coli*(S) |
| 5G | - | + | + | - | + | - | + | + | + | ++ | ++ | ++ | *E. coli*(S) |
| 6G | - | + | + | - | + | - | + | + | + | ++ | ++ | + | *E. coli*(S) |
| 7G | ++ | ++ | ++ | ++ | ++ | ++ | ++ | ++ | ++ | ++ | ++ | ++ | *K. pneumoniae*(C) |
| 8G | - | + | + | - | + | - | + | + | + | ++ | ++ | + | *E. coli*(S) |
| 9G | ++ | ++ | + | - | + | + | + | - | + | ++ | + | - | Mixed population |
| 10G | - | + | + | - | + | - | + | + | + | ++ | ++ | + | *E. coli*(S) |

| | | | | | | | | | | | | | |
|---|---|---|---|---|---|---|---|---|---|---|---|---|---|
| 11G | - | + | - | - | - | - | + | - | + | + | ++ | - | *Shigella* (S) |
| 12G | + | ++ | ++ | - | + | - | + | + | + | ++ | ++ | ++ | *E. coli*(C) |
| 13G | - | + | + | - | + | - | + | + | + | ++ | ++ | + | *E. coli* (S) |
| 14G | + | ++ | ++ | ++ | + | ++ | ++ | ++ | ++ | + | ++ | + | *K. pneumoniae*(S) |
| 15G | + | + | + | - | + | - | + | + | + | + | ++ | + | *E. coli*(S) |
| 16G | + | ++ | + | - | + | - | + | + | + | ++ | + | ++ | *E. coli*(S) |
| 17G | ++ | + | ++ | ++ | ++ | + | ++ | ++ | ++ | ++ | ++ | ++ | *K. pneumoniae*(S) |
| 18G | - | + | + | - | + | - | + | + | + | ++ | ++ | + | *E. coli*(S) |
| 19G | + | ++ | ++ | + | ++ | ++ | ++ | ++ | + | ++ | ++ | + | *K. pneumoniae*(S) |
| 20G | + | ++ | ++ | - | + | - | + | + | + | ++ | ++ | ++ | *E. coli*(C) |
| 21N | - | + | - | - | - | - | + | - | + | + | ++ | - | *Shigella* (S) |
| 22N | + | ++ | ++ | ++ | ++ | ++ | + | ++ | ++ | ++ | ++ | + | *K. pneumoniae*(S) |
| 23N | + | ++ | + | - | + | - | + | + | + | ++ | ++ | + | *E. coli*(S) |
| 24N | + | ++ | ++ | - | + | - | + | + | + | + | ++ | + | *E. coli*(S) |
| 25N | ++ | ++ | ++ | ++ | ++ | ++ | ++ | ++ | ++ | ++ | ++ | ++ | *K. pneumoniae*(S) |
| 26N | - | + | + | - | + | - | + | + | + | ++ | ++ | + | *E. coli*(S) |
| 27N | - | - | - | - | - | * | - | - | - | * | - | * | *Alcaligenes faecalis*(S) |
| 28N | + | + | ++ | - | + | - | + | + | + | ++ | + | ++ | *E. coli*(S) |
| 29N | + | ++ | ++ | - | + | - | + | + | + | ++ | ++ | ++ | *E. coli*(C) |
| 30N | - | + | + | - | + | - | + | + | + | ++ | ++ | + | *E. coli*(S) |
| 31D | + | ++ | ++ | ++ | ++ | ++ | ++ | + | ++ | ++ | ++ | ++ | *K. pneumoniae* |
| 32D | - | + | + | - | + | - | + | + | + | + | ++ | + | *E. coli*(S) |
| 33D | - | + | + | - | + | - | + | + | + | ++ | ++ | + | *E. coli*(S) |
| 34D | ++ | ++ | ++ | ++ | ++ | ++ | ++ | ++ | ++ | ++ | ++ | ++ | *K. pneumonia*(C) |
| 35D | ++ | ++ | ++ | ++ | ++ | ++ | + | ++ | ++ | ++ | ++ | + | *K. pneumonia*(S) |
| 36D | - | + | + | - | + | - | + | + | + | ++ | ++ | + | *E. coli*(S) |
| 37D | ++ | + | - | + | - | - | ++ | ++ | + | + | + | - | Mixed population |
| 38D | - | + | + | - | + | - | + | + | + | ++ | ++ | + | *E. coli*(S) |
| 39D | + | + | ++ | + | ++ | + | ++ | + | ++ | + | ++ | ++ | *K. pneumonia*(S) |
| 40D | - | - | - | - | - | * | - | - | - | * | - | * | *Alcaligenes faecalis*(S) |
| 41D | + | ++ | ++ | - | + | - | + | + | + | ++ | ++ | ++ | *E. coli*(C) |
| 42D | - | + | + | - | + | - | + | + | + | ++ | ++ | + | *E. coli*(S) |
| 43D | + | + | + | - | + | - | + | + | + | + | ++ | ++ | *E. coli*(S) |
| 44D | + | ++ | + | - | + | - | + | + | + | ++ | ++ | + | *E. coli*(S) |
| 45D | + | ++ | ++ | - | + | - | + | + | + | ++ | ++ | ++ | *E. coli*(C) |

| | | | | | | | | | | | | | |
|---|---|---|---|---|---|---|---|---|---|---|---|---|---|
| 46D | + | ++ | + | ++ | ++ | ++ | ++ | + | ++ | ++ | ++ | ++ | *K. pneumoniae*(S) |
| 47D | - | + | + | - | + | - | + | + | + | ++ | ++ | + | *E. coli*(S) |
| 48D | ++ | + | ++ | + | + | + | ++ | + | ++ | ++ | + | + | *K. pneumoniae*(S) |
| 49D | + | + | ++ | - | + | - | + | + | + | + | + | + | *E. coli*(S) |
| 50D | - | - | - | - | - | * | - | - | - | * | - | * | *Alcaligenes faecalis*(S) |
| **ZONE 2** | | | | | | | | | | | | | |
| 51A | - | + | + | - | + | - | + | + | + | ++ | ++ | + | *E. coli*(S) |
| 52A | + | ++ | ++ | - | + | - | + | + | + | ++ | ++ | ++ | *E. coli*(C) |
| 53A | - | + | + | - | + | - | + | + | + | ++ | ++ | + | *E. coli*(S) |
| 54A | ++ | ++ | ++ | ++ | ++ | ++ | ++ | ++ | ++ | ++ | ++ | ++ | *K. pneumonia*(C) |
| 55A | ++ | ++ | ++ | ++ | ++ | ++ | ++ | ++ | ++ | ++ | ++ | ++ | *K. pneumoniae*(C) |
| 56A | + | + | ++ | - | + | - | + | + | + | + | ++ | ++ | *E. coli*(S) |
| 57A | - | - | ++ | - | + | - | + | ++ | + | ++ | - | + | Mixed population |
| 58A | + | ++ | ++ | - | + | - | + | + | + | ++ | ++ | ++ | *E. coli*(C) |
| 59A | - | + | + | - | + | - | + | + | + | ++ | ++ | + | *E. coli*(S) |
| 60A | + | ++ | ++ | - | + | - | + | + | + | + | + | + | *E. coli*(S) |
| 61A | - | - | - | - | - | * | - | - | - | * | - | * | *Alcaligenes faecalis*(S) |
| 62A | ++ | ++ | ++ | ++ | ++ | ++ | ++ | ++ | ++ | ++ | ++ | ++ | *K. pneumoniae*(S) |
| 63A | - | + | + | - | + | - | + | + | + | ++ | ++ | + | *E. coli*(S) |
| 64A | - | + | + | - | + | - | + | + | + | ++ | ++ | + | *E. coli*(S) |
| 65A | + | ++ | + | ++ | ++ | + | ++ | ++ | ++ | + | ++ | + | *K. pneumoniae*(S) |
| 66A | - | + | + | - | ++ | - | + | + | ++ | ++ | ++ | - | Mixed population |
| 67A | + | + | ++ | - | + | - | + | + | + | + | ++ | ++ | *E. coli*(S) |
| 68A | + | ++ | + | - | + | - | + | + | + | ++ | ++ | ++ | *E. coli*(S) |
| 69A | - | - | - | - | - | * | - | - | - | * | - | * | *Alcaligenes faecalis*(S) |
| 70A | - | + | + | - | + | - | + | + | + | ++ | + | + | *E. coli*(S) |
| 71Al | - | + | + | - | + | - | + | + | + | + | ++ | + | *E. coli*(S) |
| 72Al | ++ | ++ | ++ | ++ | ++ | ++ | ++ | ++ | ++ | ++ | ++ | ++ | *K. pneumoniae*(C) |
| 73Al | - | + | + | - | + | - | + | + | + | ++ | ++ | + | *E. coli*(S) |
| 74Al | + | ++ | ++ | - | + | - | + | + | + | ++ | ++ | ++ | *E. coli*(C) |
| 75Al | - | + | + | - | + | - | + | + | + | ++ | + | + | *E. coli*(S) |
| 76Al | + | ++ | ++ | ++ | ++ | + | + | + | ++ | ++ | ++ | ++ | *K. pneumonia* (S) |
| 77Al | ++ | ++ | ++ | - | ++ | + | - | ++ | - | ++ | ++ | + | *Pantoea agglomerans* (C) |

| | | | | | | | | | | | | | |
|---|---|---|---|---|---|---|---|---|---|---|---|---|---|
| 78Al | - | + | + | - | + | - | + | + | + | +++ | ++ | + | *E. coli*(S) |
| 79Al | + | ++ | ++ | - | + | - | + | + | + | ++ | ++ | ++ | *E. coli*(C) |
| 80Al | - | - | - | - | - | * | - | - | - | * | - | * | *Alcaligenes faecalis*(S) |
| 81Al | + | + | + | - | + | - | + | + | + | ++ | ++ | + | *E. coli*(S) |
| 82Al | + | ++ | + | - | + | - | + | + | + | ++ | ++ | + | *E. coli*(S) |
| 83Al | ++ | ++ | ++ | ++ | ++ | ++ | ++ | ++ | ++ | ++ | ++ | ++ | *K. pneumoniae*(C) |
| 84Al | - | + | + | - | + | - | + | + | + | ++ | ++ | + | *E. coli*(S) |
| 85Al | - | + | + | - | + | - | + | + | + | ++ | ++ | + | *E. coli*(S) |
| 86Al | + | + | ++ | - | + | - | + | + | + | ++ | ++ | + | *E. coli*(S) |
| 87Al | - | - | - | - | - | * | - | * | - | * | - | * | *Alcaligenes faecalis*(S) |
| 88Al | + | ++ | + | - | + | - | + | + | + | + | ++ | ++ | *E. coli*(S) |
| 89Al | + | ++ | ++ | - | + | - | + | + | + | ++ | + | ++ | *E. coli*(S) |
| 90Al | - | ++ | + | ++ | - | ++ | + | + | ++ | ++ | ++ | + | Mixed population |
| 91H | ++ | ++ | ++ | ++ | ++ | ++ | ++ | ++ | + | ++ | ++ | ++ | *K. pneumoniae*(S) |
| 92H | + | ++ | ++ | - | + | - | + | + | + | ++ | ++ | ++ | *E. coli*(C) |
| 93H | + | + | + | - | + | - | + | + | + | ++ | ++ | ++ | *E. coli*(S) |
| 94H | - | + | + | - | + | - | + | + | + | ++ | ++ | + | *E. coli*(S) |
| 95H | + | + | ++ | - | + | - | + | + | + | ++ | ++ | + | *E. coli*(S) |
| 96H | - | + | + | - | + | - | + | + | + | ++ | ++ | + | *E. coli*(S) |
| 97H | ++ | + | ++ | + | + | ++ | + | ++ | + | + | + | + | *K. pneumonia*(S) |
| 98H | - | + | + | - | + | - | + | + | + | ++ | ++ | + | *E. coli*(S) |
| 99H | + | ++ | ++ | ++ | + | + | + | + | + | + | ++ | + | Mixed population |
| 100H | - | ++ | + | - | + | - | + | + | + | ++ | ++ | + | *E. coli*(S) |
| **ZONE 3** | | | | | | | | | | | | | |
| 101M | + | ++ | ++ | - | + | - | + | + | + | ++ | ++ | ++ | *E. coli*(C) |
| 102M | + | ++ | ++ | - | + | - | + | + | + | ++ | ++ | ++ | *E. coli*(C) |
| 103M | ++ | + | ++ | + | + | ++ | + | ++ | + | + | + | + | *K. pneumonia*(S) |
| 104M | ++ | + | ++ | ++ | + | ++ | + | + | + | + | ++ | ++ | *K. pneumoniae*(S) |
| 105M | ++ | ++ | ++ | - | ++ | + | - | ++ | - | ++ | ++ | + | *Pantoea agglomerans*(S) |
| 106M | + | + | + | - | + | - | + | + | + | ++ | ++ | ++ | *E. coli*(S) |
| 107M | - | + | + | - | + | - | + | + | + | ++ | ++ | + | *E. coli*(S) |
| 108M | ++ | ++ | ++ | ++ | ++ | ++ | ++ | ++ | ++ | ++ | ++ | ++ | *K. pneumoniae*(C) |
| 109M | - | + | + | - | + | - | + | + | + | ++ | ++ | + | *E. coli*(S) |
| 110M | + | + | + | - | + | - | + | + | + | ++ | ++ | ++ | *E. coli*(S) |

| | | | | | | | | | | | | | |
|---|---|---|---|---|---|---|---|---|---|---|---|---|---|
| 111M | + | ++ | + | ++ | ++ | ++ | ++ | + | ++ | ++ | ++ | + | *K. pneumoniae*(S) |
| 112M | - | + | + | - | + | - | + | + | + | ++ | ++ | + | *E. coli*(S) |
| 113M | - | + | ++ | - | + | - | + | + | + | ++ | ++ | + | *E. coli*(S) |
| 114M | + | ++ | + | - | + | - | + | + | + | ++ | + | ++ | *E. coli*(S) |
| 115M | + | ++ | ++ | - | + | - | + | + | + | ++ | ++ | ++ | *E. coli*(C) |
| 116M | - | + | + | - | + | - | + | + | + | ++ | ++ | + | *E. coli*(S) |
| 117M | + | ++ | + | - | + | - | + | + | + | ++ | ++ | + | *E. coli*(S) |
| 118M | ++ | ++ | - | ++ | - | + | - | + | - | ++ | - | + | Mixed population |
| 119M | - | + | + | - | + | - | + | + | + | ++ | ++ | + | *E. coli*(S) |
| 120M | - | + | + | - | + | - | + | + | + | ++ | ++ | ++ | *E. coli*(S) |
| 121B | ++ | ++ | ++ | ++ | ++ | ++ | ++ | ++ | ++ | ++ | ++ | ++ | *K. pneumoniae*(C) |
| 122B | - | + | + | - | + | - | + | + | + | ++ | ++ | ++ | *E. coli*(S) |
| 123B | + | + | ++ | - | + | - | + | + | + | ++ | ++ | + | *E. coli*(S) |
| 124B | + | ++ | + | - | + | - | + | + | + | ++ | ++ | + | *E. coli*(S) |
| 125B | - | + | + | - | + | - | + | + | + | ++ | ++ | ++ | *E. coli*(S) |
| 126B | - | + | + | - | + | - | + | + | + | ++ | ++ | + | *E. coli*(S) |
| 127B | ++ | ++ | + | ++ | + | + | + | + | ++ | ++ | ++ | + | *K. pneumoniae*(S) |
| 128B | ++ | ++ | + | ++ | ++ | ++ | ++ | ++ | + | ++ | ++ | ++ | *K. pneumoniae*(S) |
| 129B | - | + | + | - | + | - | ++ | + | + | ++ | ++ | - | Mixed population |
| 130B | ++ | ++ | ++ | ++ | ++ | ++ | ++ | ++ | ++ | ++ | ++ | ++ | *K. pneumoniae*(S) |
| 131B | - | + | + | - | + | - | + | + | + | + | ++ | + | *E. coli*(S) |
| 132B | + | ++ | + | ++ | ++ | ++ | ++ | ++ | + | ++ | ++ | ++ | *K. pneumoniae*(S) |
| 133B | + | ++ | ++ | - | + | - | + | + | + | ++ | ++ | ++ | *E. coli*(C) |
| 134B | + | ++ | ++ | - | + | - | + | + | + | ++ | ++ | ++ | *E. coli*(C) |
| 135B | - | + | + | - | + | - | + | + | + | ++ | ++ | + | *E. coli*(S) |
| 136B | ++ | ++ | ++ | ++ | ++ | ++ | ++ | ++ | ++ | ++ | ++ | ++ | *K. pneumoniae*(C) |
| 137B | - | + | + | - | + | - | + | + | + | ++ | ++ | + | *E. coli*(S) |
| 138B | + | ++ | ++ | + | ++ | + | + | + | + | + | + | + | *K. pneumoniae*(S) |
| 139B | + | ++ | ++ | - | + | - | + | + | + | ++ | ++ | ++ | *E. coli*(C) |
| 140B | - | + | + | - | + | - | + | + | + | ++ | ++ | + | *E. coli*(S) |
| 141Bj | ++ | ++ | + | ++ | ++ | ++ | ++ | ++ | ++ | ++ | ++ | + | *K. pneumoniae*(S) |
| 142Bj | - | ++ | ++ | - | ++ | - | ++ | - | - | - | ++ | - | *S. enteritidis* (S) |
| 143Bj | + | ++ | + | - | + | - | + | + | + | ++ | ++ | + | *E. coli*(S) |
| 144Bj | + | + | ++ | - | + | - | + | + | + | ++ | ++ | - | *E. coli*(S) |
| 145Bj | ++ | ++ | ++ | ++ | ++ | ++ | ++ | ++ | ++ | ++ | ++ | ++ | *K. pneumoniae*(S) |
| 146Bj | - | + | + | - | + | - | + | + | + | ++ | ++ | + | *E. coli*(S) |
| 147Bj | - | + | + | - | + | - | ++ | - | + | + | ++ | + | Mixed population |

| | | | | | | | | | | | | | |
|---|---|---|---|---|---|---|---|---|---|---|---|---|---|
| 148Bj | + | + | ++ | - | + | - | + | + | + | ++ | ++ | - | *E. coli*(S) |
| 149Bj | - | + | + | - | + | - | + | + | + | ++ | ++ | + | *E. coli*(S) |
| 150Bj | + | ++ | ++ | - | + | - | + | + | + | ++ | ++ | ++ | *E. coli*(C) |

**Note:** ++ Positive (more than 90%)　+ Positive (11-89%)　- Negative (more than 90%)　*Data not available

TDA: Phenylalanine Deaminase Test　ONPG: O-Nitrophenyl-β-D-galactopyranoside Test

Colouring shows highly resistant strains identified by VITEK
